# A novel lncRNA FAM151B-DT regulates autophagy and degradation of aggregation prone proteins

**DOI:** 10.1101/2025.01.22.25320997

**Authors:** Arun Renganathan, Miguel A. Minaya, Matthew Broder, Isabel Alfradique-Dunham, Michelle Moritz, Reshma Bhagat, Jacob Marsh, Anthony Verbeck, Grant Galasso, Emma Starr, David A. Agard, Carlos Cruchaga, Celeste M. Karch

## Abstract

Neurodegenerative diseases share common features of protein aggregation along with other pleiotropic traits, including shifts in transcriptional patterns, neuroinflammation, disruptions in synaptic signaling, mitochondrial dysfunction, oxidative stress, and impaired clearance mechanisms like autophagy. However, key regulators of these pleotropic traits have yet to be identified. Here, we discovered a novel long non-coding RNA (lncRNA), *FAM151B-DT*, that is reduced in a stem cell model of frontotemporal dementia with tau inclusions (FTLD-tau) and in brains from FTLD-tau, progressive supranuclear palsy, Alzheimer’s disease, and Parkinson’s disease patients. We show that silencing *FAM151B-DT in vitro* is sufficient to enhance tau aggregation. To begin to understand the mechanism by which *FAM151B-DT* mediates tau aggregation and contributes to several neurodegenerative diseases, we deeply characterized this novel lncRNA and found that *FAM151B-DT* resides in the cytoplasm where it interacts with tau, α-synuclein, HSC70, and other proteins enriched in protein homeostasis. When silenced, *FAM151B-DT* blocks autophagy, leading to the accumulation of tau and α-synuclein. Importantly, we discovered that increasing *FAM151B-DT* expression is sufficient to promote autophagic flux, reduce phospho-tau and α-synuclein, and reduce tau aggregation. Overall, these findings pave the way for further exploration of *FAM151B-DT* as a promising molecular target for several neurodegenerative diseases.

## Introduction

Tauopathies are complex disorders characterized by a range of molecular, cellular, and physiological changes beyond tau inclusions ^1, 2, 3^. Tauopathy etiology is pleiotropic, encompassing a cascade of events such as shifts in transcriptional patterns, translational stress, perturbations in the function of RNA binding proteins, inflammation, disruptions in synaptic functioning, neurotransmitter imbalance, mitochondrial dysfunction, oxidative stress, and impaired cellular clearance mechanisms like autophagy ^4, 5, 6, 7^. Yet, the upstream molecular drivers of these events remain elusive.

A substantial fraction of the human genome is transcribed into RNAs that do not encode proteins. This non-coding genome constitutes over 98% of the total human genome and was once referred to as “junk DNA”. Next generation sequencing technologies have revealed that the non-coding genome plays essential pleiotropic roles in regulation and is involved in various cellular processes ^8, 9^. Long non-coding RNAs (lncRNAs), transcripts >200bp, play crucial roles in regulating gene expression at multiple levels, including chromatin remodeling, transcriptional, and post-translational regulation ^10^. LncRNAs also function as molecular scaffolds, decoys, or guides, influencing the assembly of protein complexes and the localization of regulatory molecules ^10^. LncRNAs are an attractive drug target in disease due to their tissue-specific expression patterns, which result in fewer undesired, off-target toxic effects ^8, 9, 10^. Yet, in the context of tauopathies, few studies have systematically evaluated the impact of lncRNAs on disease.

To begin to systematically evaluate the impact of lncRNAs in tauopathies, we have recently applied transcriptomics, cellular, and molecular biology approaches to frontotemporal lobar dementia (FTLD) caused by mutations in the *MAPT* gene (FTLD-tau) ^11^. Using stem cell derived neurons from patients carrying a *MAPT* p.P301L, IVS10+16, or p.R406W mutation, and CRISPR-corrected isogenic controls, we identified 15 lncRNAs that were commonly differentially expressed across the three *MAPT* mutations ^11^. These lncRNAs interact with RNA binding proteins involved in FTD, including FUS and TDP43, and regulate stress granule formation ^11^. Here, we build upon our prior work and describe a novel lncRNA, *FAM151B-DT*. We show that *FAM151B-DT* expression is reduced in *MAPT* mutant neurons and brains from tauopathy patients. We find that *FAM151B-DT* mitigates tau aggregation by acting as a scaffold for HSC70 and tau to promote clearance through autophagic pathways. Together, our work suggests that novel lncRNA *FAM151B-DT* may serve as a novel therapeutic target for tauopathies.

## Results

### LncRNA FAM151B-DT is reduced in tauopathies

We have previously shown that iPSC-derived neurons expressing *MAPT* mutations (IVS10+16, P301L, and R406W) are sufficient to alter expression of 15 lncRNAs (**Figure 1A**)^11^. While these included a number of well-annotated lncRNAs, such as *NORAD*, *SOX9-AS1*, *GAPBB1-AS1*, and *SNHG8*, we also identified several poorly annotated lncRNAs with unknown function^11^. Among these 15 differentially expressed lncRNAs, *AC008771.1* (*FAM151B-DT*) was significantly reduced in *MAPT* IVS10+16, P301L, and R406W iPSC-derived neurons compared with isogenic controls (**Figure 1B**; **Supplemental Table 1**).

**Figure 1.**
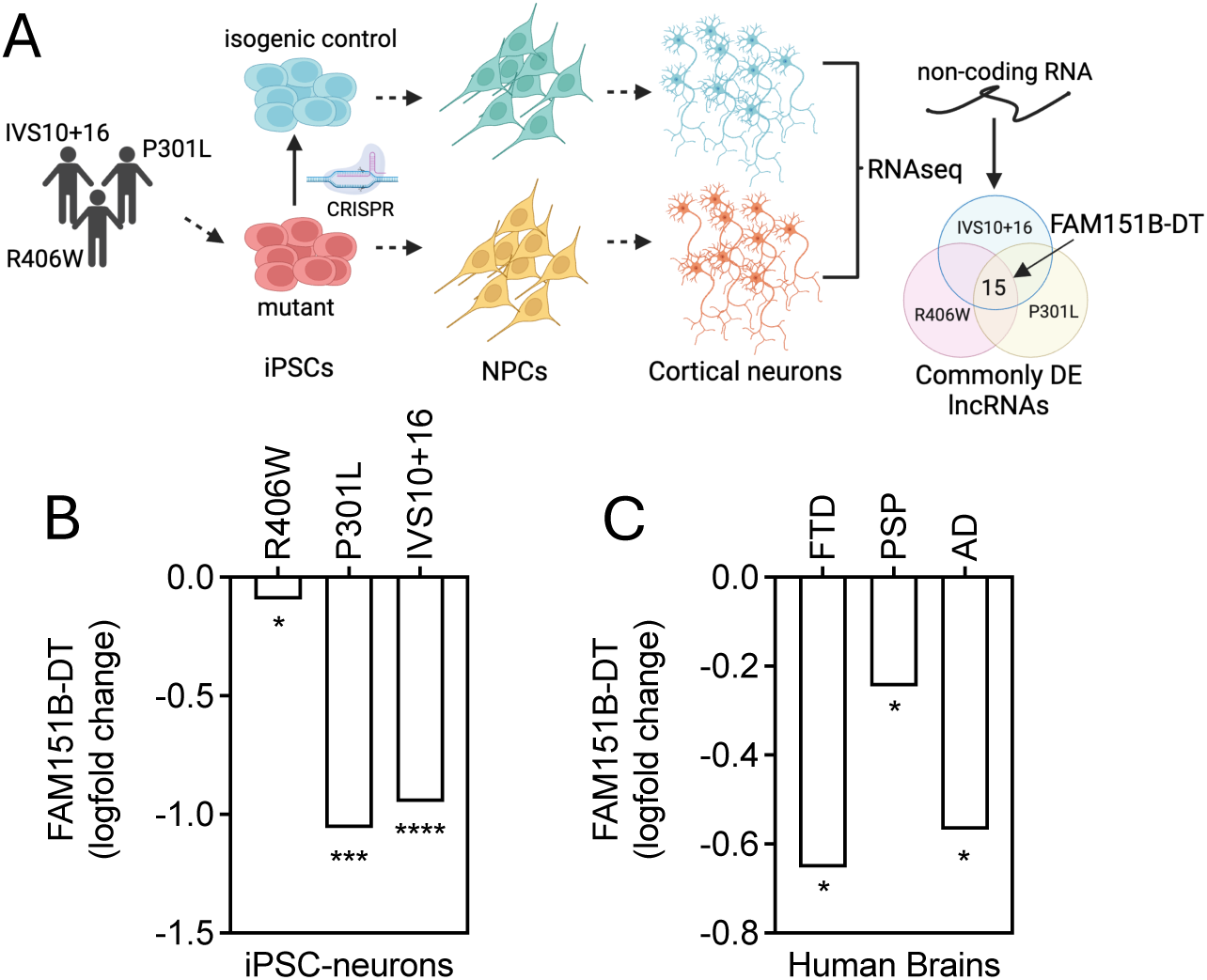
*FAM151B-DT* expression is significantly reduced in *MAPT* mutant neurons and tauopathy brains. A. Schematic representation of the discovery of lncRNAs that are differentially expressed (DE) in *MAPT* mutant iPSC-neurons compared with isogenic controls. *FAM151B-DT* was among the 15 common dysregulated lncRNAs in *MAPT* R406W, P301L, and IVS10+16 neurons ^11^. B. *FAM151B-DT* is significantly reduced in *MAPT* R406W, P301L, and IVS10+16 neurons compared with isogenic controls. Graph represents Log_2_FC. Data is representative of three biological replicates. C. FAM151B-DT is significantly reduced in human FTD with tau inclusions, AD, PSP brains compared with control brain tissues. Graph represents Log_2_FC. *, p≤0.05; ***, p<0.001; ****, p<0.0001.

We next sought to determine whether *FAM151B-DT* was altered in brains from tauopathy patients. We found that *FAM151B-DT* was significantly reduced in brains from *MAPT* mutation carriers with FTLD-tau pathology, sporadic primary tauopathy progressive supranuclear palsy (PSP), and secondary tauopathy Alzheimer’s disease (AD; **Figure 1C**; **Supplemental Table 1**). Within these cohorts, *FAM151B-DT* was robustly expressed among the lncRNAs that passed QC (see Methods; **Supplemental Figure 1**). Together, these findings illustrate that reduced *FAM151B-DT* is a common feature in *MAPT* mutant neurons *in vitro* and in the presence of tau pathology *in vivo*.

### FAM151B-DT modulates tau seeding

Given our discovery that *FAM151B-DT* is reduced in *MAPT* mutant neurons and tauopathy brains, we sought to determine whether *FAM151B-DT* impacts the propensity for tau to seed new aggregates. To do this, we used a monoclonal biosensor cell model whereby a fragment of tau containing the microtubule binding domain (MTBR) is engineered with the *MAPT* P301S mutation and tagged with a CFP or YFP ^12^. Addition of tau fibrils to these tau biosensor cells promotes the two tagged forms of tau to aggregate and generate a FRET signal^12^.

To determine how reduction of *FAM151B-DT* impacts tau seeding, tau biosensor cells were transiently transfected with siRNA directed to *FAM151B-DT* or a scrambled control for 24 hours (**Figure 2A**). Tau biosensor cells were then treated with seed competent tau (Tau-441) for 48 hours (**Figure 2A**). siRNAs specific to *FAM151B-DT* produced approximately 50% reduction in endogenous *FAM151B-DT* expression in the tau biosensor cells (**Figure 2B**). Silencing of *FAM151B-DT* resulted in a statistically significant increase in tau seeding (**Figure 2C**; **Supplemental Figure 2A**). Thus, decreasing *FAM151B-DT* in a manner consistent with observed changes in *MAPT* mutant neurons and tauopathy brains promotes tau aggregation *in vitro*.

**Figure 2.**
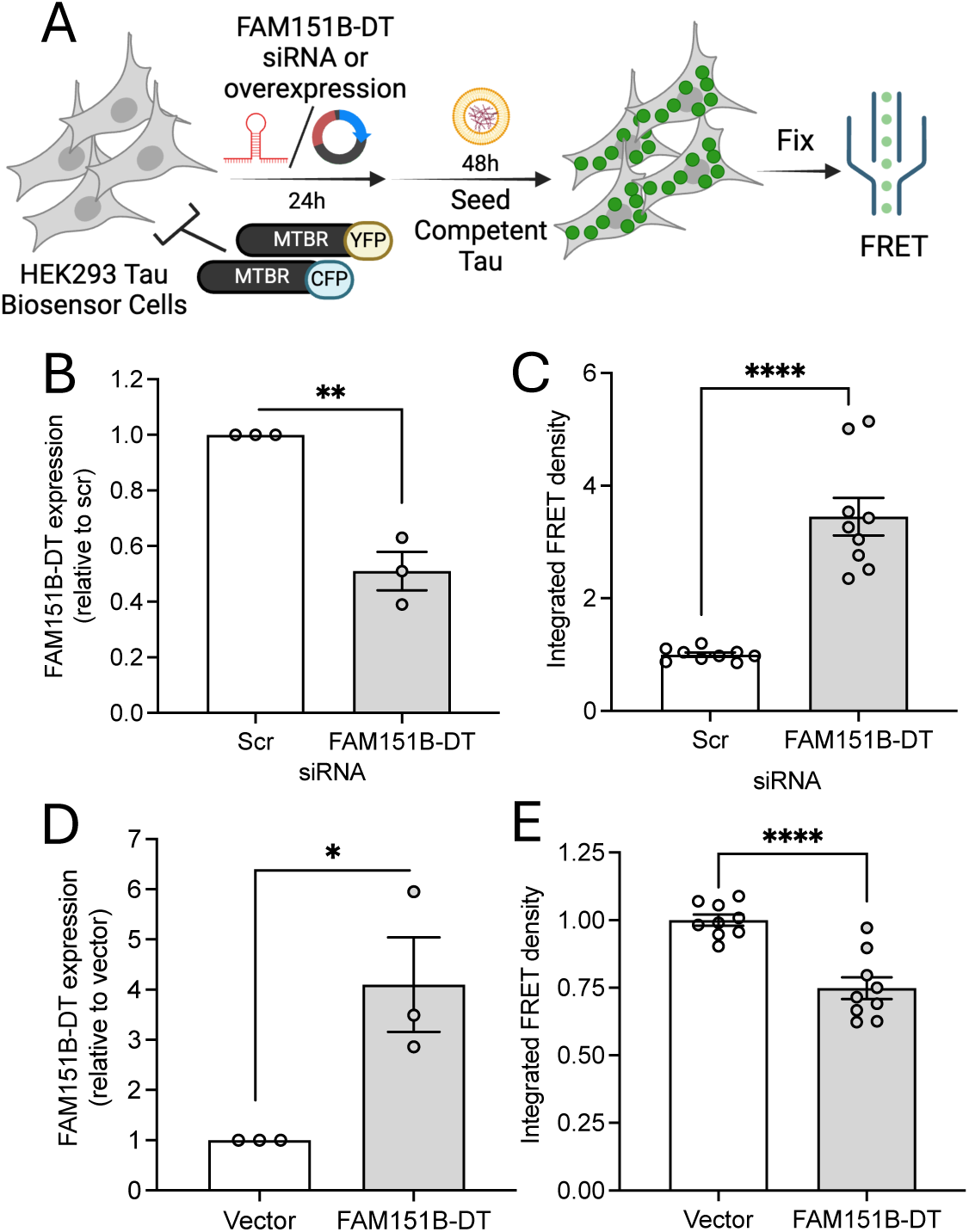
*FAM151B-DT* expression regulates tau seeding. A. Schematic representation of the FRET-based tau seeding assay. B. qPCR for *FAM151B-DT* reveals significant reduction of *FAM151B-DT* levels after siRNA treatment in tau biosensor cells. C. Silencing *FAM151B-DT* significantly increased integrated FRET density compared to control siRNA-treated tau biosensor cells. Quantification of the FRET signal, normalized to controls. Error bars represent SEM, n = 50,000 cells per experiment. D. qPCR for *FAM151B-DT* reveals significant increase in *FAM151B-DT* levels upon overexpression in tau biosensor cells. E. *FAM151B-DT* overexpression significantly decreased the integrated FRET density compared to ctrl-vector-treated tau-biosensor cells. Quantification of the FRET signal, normalized to controls. Error bars represent SEM, n = 50,000 cells per experiment. Data is representative of three biological replicates. Student’s t-test, *, p≤0.05; **, p<0.001; ****, p<0.0001.

We next sought to determine how overexpression of *FAM151B-DT* impacts tau seeding in the tau biosensor cells (**Figure 2A**). Plasmids containing an empty vector or *FAM151B-DT* were transiently transfected in the tau biosensor cells for 24 hours, and the resulting cells were treated with tau seeds (Tau-441) for 48 hours (**Figure 2A**). In the context of ∼4-fold overexpression of *FAM151B-DT* (**Figure 2D**), tau seeding was significantly reduced (**Figure 2E**; **Supplemental Figure 2B**). Thus, increasing *FAM151B-DT* overexpression rescues tau seeding propensity. Together, this work illustrates that *FAM151B-DT* modulation regulates tau seeding.

### Characterizing the novel lncRNA FAM151B-DT

To define the mechanism by which *FAM151B-DT* regulates tau seeding and contributes to tauopathy, we began to carefully characterize this novel lncRNA. At the time of our discovery that *AC008771.1* (*FAM151B-DT*) was significantly reduced in *MAPT* mutant neurons and tauopathy brains, little information was available regarding the features of this lncRNA. UCSC-PhastCons pairwise conservation analysis reveals that *FAM151B-DT* is poorly expressed across 100 vertebrates and 13 mammals (**Supplemental Figure 3**). LncRNAs tend to exhibit more restricted expression patterns than mRNAs ^13, 14^; however, *FAM151B-DT* is moderately expressed across tissues in the body (**Supplemental Figure 4A**). While *FAM151B-DT* is expressed across the cortex, brain stem, and spinal cord (**Supplemental 4B**), its highest expression occurs in regions largely spared of tau pathology (e.g. cerebellum hemispheres; **Supplemental 4B**). Within the brain, *FAM151B-DT* is most highly expressed in neurons, but it remains detectable across cell-types, including astrocytes, oligodendrocytes, oligodendrocyte precursors, microglia, and endothelial cells (**Supplemental Figure 4C**). Together, these findings illustrate that *FAM151B-DT* is a poorly conserved lncRNA that is widely expressed in the CNS.

LncRNAs can act as cis-regulatory elements by modulating the expression of neighboring genes through proximity-based mechanisms. *FAM151B-DT* is located on chromosome 5q12.1 (chr5:80411231-80487968) where it is transcribed from the negative strand in a divergent orientation relative to the adjacent protein-coding gene, *FAM151B*, with which it shares a promoter region (**Supplemental Figure 5**). *ZFYVE16*, a zinc finger-related protein, is transcribed from the positive strand in close proximity to *FAM151B-DT* (**Supplemental Figure 5A**). Such genomic arrangements are often associated with transcriptional cross-regulation ^15^. To explore whether *FAM151B-DT* exerts cis-regulatory effects, we silenced FAM151B-DT in SH-SY5Y cells and analyzed the expression levels of *FAM151B* and *ZFYVE16*. With nearly 80% knockdown of endogenous *FAM151B-DT* (**Supplemental Figure 5B**), we observed no significant changes in the transcript levels of *FAM151B* or *ZFYVE16* (**Supplemental Figure 5C-D**). Consistent with this finding, *FAM151B* and *ZFYVE16* expression was variable across *MAPT* mutant neurons (**Supplemental Figure 5E-F**) and unchanged in tauopathy brains (**Supplemental Figure 5G**). Thus, *FAM151B-DT* does not act as a cis-regulatory element in SH-SY5Y, iPSC-derived neurons or human brains.

To further explore potential functions of *FAM151B-DT*, we evaluated subcellular localization of *FAM151B-DT*. LncRNAs can play crucial roles in regulating gene expression at multiple levels including chromatin remodeling, transcriptional, and post-translational regulation, as well as function as molecular scaffolds, decoys, or guides, influencing the assembly of protein complexes and the localization of regulatory molecules ^13^. The subcellular localization of a given lncRNA can point to one or more of these functions. To define *FAM151B-DT* localization, we performed biochemical fractionation coupled with RNA quantification in iPSC-derived neurons (**Figure 3A**) and SH-SY5Y cells (**Figure 3B**). The nuclear enriched lncRNA, *MALAT1* ^16^, and the cytoplasmic enriched lncRNA, *TUG1* ^17^, were included as controls (**Figure 3A-B**). We found that *FAM151B-DT* is enriched in the cytoplasm in iPSC-derived neurons and SH-SY5Y cells, consistent with the pattern observed for *TUG1* (**Figure 3A-B**). *FAM151B-DT*’s enrichment in the cytoplasm is also consistent with our prior observations that *FAM151B-DT* does not act as a cis-regulatory element.

**Figure 3.**
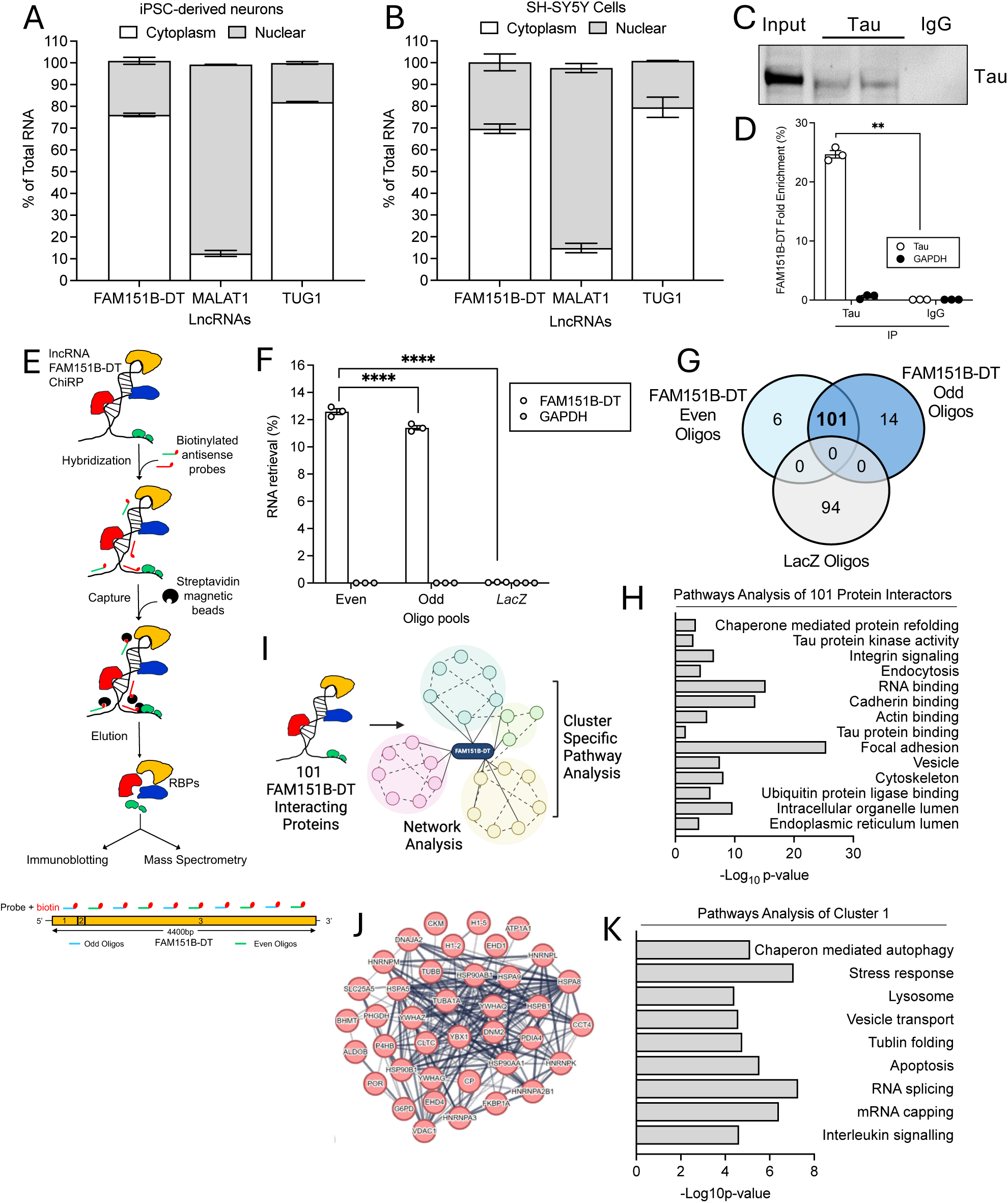
Proteins interacting with *FAM151B-DT*. A-B. Nuclear and cytosolic isolation of RNAs to define the localization of *FAM151B-DT* relative to well characterized lncRNAs (nuclear localized *MALAT1* and cytosolic *TUG1*). A. iPSC-derived neurons. B. SH-SY5Y cells. C-D. Tau interacts with *FAM151B-DT*. C. Immunoprecipitation of tau (Tau5) in SH-SY5Y cells. D. RNA immunoprecipitation (RIP)-qPCR validation of the tau-bound RNA fraction shows robust *FAM151B-DT* expression. *GAPDH* included as a negative control. Data is representative of three biological replicates. Two-way ANOVA, ****, p<0.0001. E. Comprehensive identification of RNA binding proteins (ChIRP) workflow. Schematic representation includes the positions of even and odd probes along the entire *FAM151B-DT* RNA sequence. F. ChIRP-qPCR reveals enrichment of *FAM151B-DT*, compared to the housekeeping transcript *GAPDH*. One way ANOVA, ****, p<0.0001. G. ChIRP-mass spectrometry hits of proteins retrieved by even and odd oligos specific for *FAM151B-DT* compared with non-targeting *LacZ* control oligos. 215 total proteins were identified across the three samples using a protein probability ≥99%; ≥2 peptides mapped; and ≥95% peptide probability (coverage). H. GO enrichment analysis of the *FAM151B-DT* proteome (101 proteins) in SH-SY5Y cells. I. Schematic of string network analysis of *FAM151B-DT* proteome reveals multiple clusters (see Supplemental Figure 5). J. Visualizing cluster 1. Protein-protein interaction enrichment p-value = 1.0e^−16^. K. Top 10 most significant pathways in cluster 1 identified by KEGG-Pathway enrichment analysis.

A finding that *FAM151B-DT* is expressed in the cytoplasm could point to a role for *FAM151B-DT* serving as a scaffold for tau, which is also located in the cytoplasm. To determine whether *FAM151B-DT* and tau interact, we performed an immunoprecipitation for tau (antibody: Tau5) in SH-SY5Y cells (**Figure 3C**), a neuroblastoma cell line used as an immortalized surrogate for neurons. SH-SY5Y cells can be readily expanded and express endogenous tau and FAM151B-DT. RNA immunoprecipitation (RIP)-qPCR validation of the tau-bound RNA fraction revealed *FAM151B-DT* (**Figure 3D**). Thus, we show for the first time that *FAM151B-DT* interacts with tau, which supports several potential mechanisms by which this lncRNA regulates tau seeding.

### FAM151B-DT interacts with a broad protein network involved in protein homeostasis

In the cytoplasm, *FAM151B-DT* may interact with a number of proteins. To begin to define the protein interactome for *FAM151B-DT*, we performed Comprehensive identification of RNA-binding proteins (ChIRP) in SH-SY5Y cells, a highly specific technique designed to isolate RNA-bound proteins (**Figure 3E**) ^18^. Oligonucleotide probes were designed across the full-length *FAM151B-DT* sequence and labeled “odd” or “even” (**Figure 3E**; **Supplemental Table 2**), ensuring efficient retrieval of the lncRNA. ChIRP-qPCR analysis confirmed strong enrichment of *FAM151B-DT* compared to the non-targeting *GAPDH* transcript (**Figure 3F**). After cross-linking and immunoprecipitation, the *FAM151B-DT* protein interactome was mapped using mass spectrometry (ChIRP-MS). After correcting for non-specific protein interactors (those isolated in the LacZ control), we identified 121 high-confidence protein interactors of *FAM151B-DT* across both probe sets (**Figure 3G**; **Supplemental Table 3**). Identified proteins met a stringent criteria of protein probability >99%, with at least two peptides required, and >95% peptide coverage. Among the 121 *FAM151B-DT* protein interactors, 101 were shared across the “odd” and “even” probes; so, we focused on these proteins for subsequent analyses (**Figure 3G**; **Supplemental Table 3**).

To continue to understand the function of *FAM151B-DT*, we began to characterize the 101 *FAM151B-DT* protein interactors. Gene Ontology (GO) enrichment analysis of the *FAM151B-DT* interactome revealed pathways involved in tau and the cytoskeleton, chaperone mediated autophagy, and endolysosomal pathways (**Figure 3H**; **Supplemental Table 4**). To better understand the degree to which these 101 proteins interact with one another to form functional protein networks, we performed STRING network analysis (**Figure 3I**; **Supplemental Figure 6**; **Supplemental Table 5**). Among the 4 major network clusters that arose from the STRING analysis, Cluster 1 contained the most *FAM151B-DT* protein interactors (**Figure 3J**; **Supplemental Table 5**). Cluster 1 was enriched in proteins involved in chaperone mediated autophagy, stress response, endolysosomal function, and apoptosis, among others (**Figure 3K**; **Supplemental Table 6**). Many of these *FAM151B-DT* interacting proteins were also differentially expressed in *MAPT* mutant neurons (**Supplemental Figure 7A**; **Supplemental Table 7**) and tauopathy brains (**Supplemental Figure 7B**; **Supplemental Table 8**). Our novel map of *FAM151B-DT* interactors point to proteins involved in protein homeostasis and disrupted in neurodegenerative processes.

### FAM151B-DT and tau share interacting partners that are disrupted in MAPT mutant neurons and human brains

Tau is a microtubule interacting protein and interacts with a broad set of proteins across functional classes ^19^. Thus, we sought to determine whether the *FAM151B-DT* interactome overlaps with the tau interactome (**Figure 4A**). We queried publicly available proteomics datasets derived from iPSC-derived neurons expressing *MAPT* WT, P301L, and V337M ^20^, soluble tau from AD brains ^21, 22, 23^, and neurofibrillary tangles (NFT) from AD brains ^24^ (**Figure 4A**). Within iPSC-derived neurons, 72 of the 101 *FAM151B-DT* interactors (71%) were also found to interact with tau (**Supplemental Table 9**). Notably, 19 proteins among the *FAM151B-DT* proteome were also found to interact with tau in iPSC-derived neurons, soluble tau in AD brains, and NFT in AD brains (**Figure 4B; Supplemental Table 9**). The proteins that interact with *FAM151B-DT* and tau are enriched in pathways involved in tau and the cytoskeleton, chaperone mediated autophagy, and endolysosomal pathways (**Figure 4C**; **Supplemental Table 10**). Several of the 19 proteins were also differentially expressed in *MAPT* mutant iPSC-derived neurons and tauopathy brains (**Figure 4D**; **Supplemental Table 7-8**). These include a number of genes encoding heat shock proteins and proteins involved in autophagy. One gene, *HSPA8*, was significantly reduced in *MAPT* P301L, IVS10+16, and R406W neurons (compared to isogenic controls), as well as FTLD-tau, PSP, and AD brains (compared to neuropathology free controls; **Figure 4D**).

**Figure 4.**
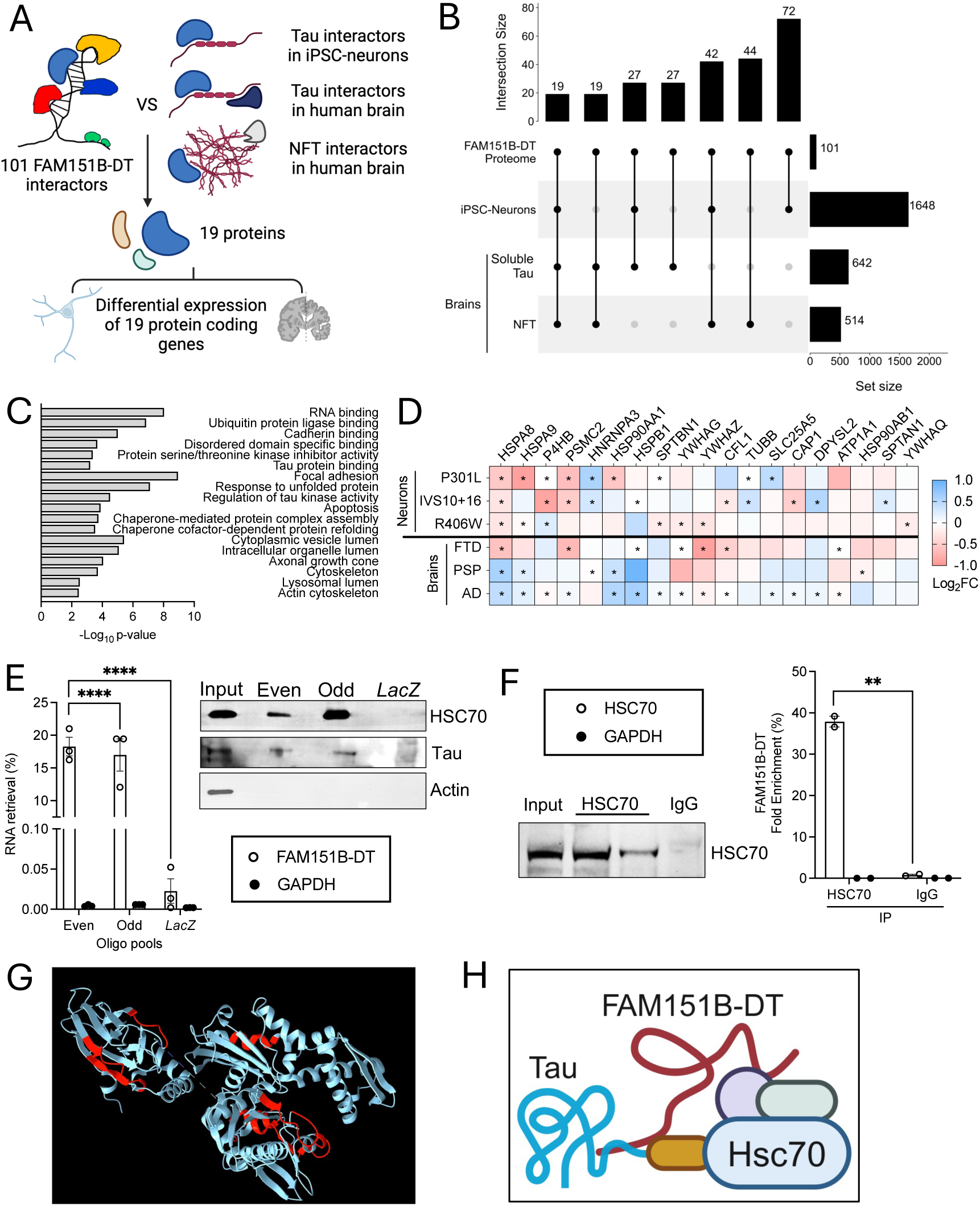
*FAM151B-DT* and tau share interacting partners that are disrupted in *MAPT* mutant neurons and human brains. A. Schematic illustrating workflow. The 101 *FAM151B-DT* interactors were compared with tau interactors defined in iPSC-neurons, human brains, and neurofibrillary tangles (NFT). The common set of interactors were then evaluated for differential expression at the RNA level in *MAPT* mutant iPSC-neurons and FTLD-tau brains. B. Upset plot for *FAM151B-DT* interactors that are also reported to interact with tau in human iPSC-derived neurons from *MAPT* WT, P301L, and V337M ^20^, soluble, total tau from human post-mortem brain tissues from AD patients ^21, 22, 23^, and NFT from AD patient brains ^24^. C. GO enrichment analysis of the *FAM151B-DT* interactome overlapped with tau and NFT interactomes (n=19). D. Heatmap representing differential expression of *FAM151B-DT*/tau shared interactome in *MAPT* mutant neurons and human brains from FTD with tau inclusions, PSP, and AD patients. *, p≤0.05. E. ChIRP-qPCR validation of FAM151B-DT pull-down using biotinylated DNA oligos. *GAPDH* was included as a negative control. Data are represented as mean ± SEM from three replicates. One-way ANOVA, ****, p< 0.0001. Right panel, ChIRP-immunoblot of *FAM151B-DT* pull-down fractions probed with HSC70 and tau (Tau5) antibodies. F. Immunoprecipitation of HSC70 in SH-SY5Y cells. Right panel, RIP-qPCR validation of HSC70-bound RNA fractions illustrates robust *FAM151B-DT* expression. *GAPDH* included as a negative control. Data are representative of three biological replicates. Two-way ANOVA, ** p < 0.01. G. Crystal structure HSC70 (PDB: 4FL9) with *FAM151B-DT* interaction sites annotated in red. H. Schematic of the *FAM151B-DT*-HSC70-tau complex.

### FAM151B-DT interacts with the HSC70/tau Complex

*HSPA8* encodes HSC70, a chaperone that functions to promote proper folding and stabilization of proteins and to facilitate degradation via the lysosome. HSC70 (e.g. *HSPA8*) interacts with both *FAM151B-DT* and tau ^25^. To further validate our ChIRP-MS (for FAM151B-DT interactors) and the publicly available proteomics datasets (for tau interactors), we performed ChIRP assays in SH-SY5Y cells. Biotinylated *FAM151B-DT* oligonucleotides efficiently pulled down endogenous HSC70 and tau proteins, which were detected by immunoblot analysis (**Figure 4E**). Protein immunoprecipitation assays with an HSC70 antibody in SH-SY5Y cells further verified that *FAM151B-DT* interacts with HSC70, as qPCR detection of the lncRNA in the pulled-down fraction showed significant enrichment (**Figure 4F**). ChIRP-MS data allowed us to map the sites on HSC70 where the interaction with *FAM151B-DT* occurs (**Figure 4G**). These complementary techniques demonstrate that *FAM151B-DT* interacts with tau and HSC70 (**Figure 4H**).

### FAM151B-DT modulates autophagic pathways

Given our findings that *FAM151B-DT* interacts with a number of proteins involved in protein homeostasis, including HSC70, we sought to determine whether *FAM151B-DT* can directly regulate these protein homeostasis pathways. *FAM151B-DT* silencing in SH-SY5Y cells did not alter HSC70 protein levels (**Figure 5A-B**). Next, we evaluated whether *FAM151B-DT* silencing impacts other key components of the autophagy/lysosome pathway. Levels of LAMP1, a marker of lysosomal integrity and autophagosome-lysosome fusion, remained unchanged (**Figure 5A-B**). Interestingly, *FAM151B-DT* silencing resulted in a significant increase in LAMP2A protein levels (**Figure 5A-B**), a key receptor on the lysosome that promotes autophagic flux ^26^. *FAM151B-DT* silencing also resulted in a significant increase in the ratio of LC3II/I and p62 (**Figure 5A-B**). The observed increase in LC3B-II/I ratio, a marker of autophagosome formation, suggests silencing *FAM151B-DT* enhances autophagy initiation. Yet, the concomitant increase in p62, cargo degraded via autophagy, suggests that autophagic degradation is inefficient. Consistent with a finding that general cargo accumulates when *FAM151B-DT* is silenced, we found that ptau and total tau were also significantly elevated (**Figure 5A-B**). Together, we demonstrate that *FAM151B-DT* silencing promotes unproductive autophagy that leads to an increase in ptau, total tau, and general cargo (e.g. p62). NFTs in human tauopathy brains are marked by the accumulation of hyperphosphorylated tau protein aggregates that are p62 positive ^27^. Together, these findings support a mechanism by which *FAM151B-DT* drives tau seeding.

**Figure 5.**
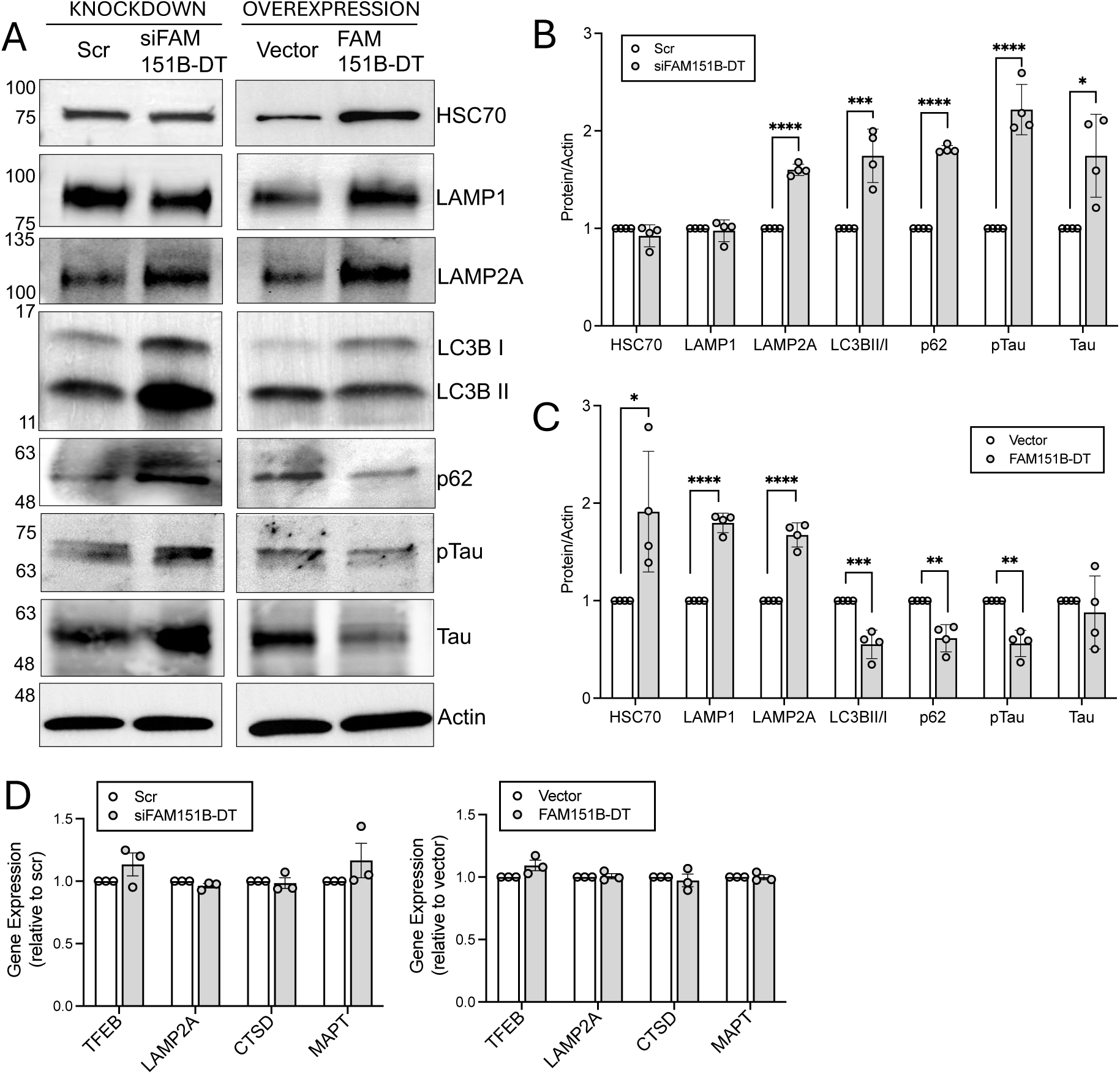
*FAM151B-DT* modulates autophagy and tau. SH-SY5Y cells were transiently transfected with scrambled (scr) or *siFAM151B-DT* siRNAs and control vector or *FAM151B-DT* overexpression plasmid. A. Representative immunoblots of SH-SY5Y cell lines detecting HSC70, lysosome (LAMP1, LAMP2A) and autophagy (LC3 and p62) markers, and tau (AT8, ptau and Tau5, total tau). B-C. Quantification of western blot data from (A). B. Quantification of *FAM151B-DT* silencing. White bar, scr. Gray bar, siFAM151B-DT. C. Quantification of *FAM151B-DT* overexpression. White bar, vector. Gray bar, *FAM151B-DT*. D-E. RNA expression of *TFEB*, *LAMP2A*, *CTSD* (Cathepsin D) and *MAPT*. D. Quantification of *FAM151B-DT* silencing. White bar, scr. Gray bar, *siFAM151B-DT*. E. Quantification of *FAM151B-DT* overexpression. White bar, vector. Gray bar, FAM151B-DT. Data are presented as mean ± SEM from four independent experiments. Student’s t-test, *, p<0.05; **, p<0.01; ***, p<0.001, and ****, p<0.0001.

To understand how *FAM151B-DT* overexpression may promote a cellular environment that reduces tau seeding, we overexpressed *FAM151B-DT* in SH-SY5Y cells. We found that when *FAM151B-DT* was elevated, HSC70, LAMP1, and LAMP2A proteins were also significantly elevated (**Figure 5A and 5C**). Overexpression of *FAM151B-DT* also resulted in a significant reduction of LC3II/I and p62 (**Figure 5A and 5C**). Together, these changes point to *FAM151B-DT* overexpression promoting an increase in lysosomes and more efficient autophagic flux. Consistent with this finding, *FAM151B-DT* overexpression resulted in a significant decrease in ptau without altering total tau (**Figure 5A and 5C**). Macroautophagy is the major pathway by which ptau is degraded ^28, 29, 30^. Thus, we propose that *FAM151B-DT* overexpression promotes degradation of aggregation prone forms of tau by macroautophagy which may ultimately reduce tau seeding.

Given our discovery that regulating *FAM151B-DT* expression impacts lysosomal/autophagic pathways, we sought to determine whether *FAM151B-DT* acts more broadly on regulators of lysosomal biogenesis and protein degradation. We evaluated mRNA levels of *TFEB* and *CTSD* (encoding Cathepsin D), key regulators of lysosomal biogenesis and function ^31, 32^. TFEB enhances Cathepsin D expression, contributing to lysosomal proteolysis and cellular clearance mechanisms ^33^. However, neither *FAM151B-DT* silencing (**Figure 5D**) nor overexpression (**Figure 5E**) produced a significant change in *TFEB*, *LAMP2A*, or *Cathepsin D* (*CTSD*). We also found that *MAPT* mRNA levels were unchanged (**Figure 5D-E**). Thus, *FAM151B-DT* likely functions at the protein level to modify autophagy and alter tau accumulation.

### FAM151B-DT interacts with autophagy substrates implicated in Parkinson’s disease

Efficient autophagy is responsible for the degradation of many proteins known to accumulate in neurodegenerative diseases ^7, 34, 35^. Given our findings that *FAM151B-DT* plays a role in modulating autophagy, we asked whether this lncRNA may act on other proteins implicated in neurodegeneration. In a subset of autophagy, the chaperone HSC70 recognizes and binds to proteins with a specific peptide motif (KFERQ) and guides the protein to the lysosome for degradation ^36, 37^. HSC70 recognizes proteins, like tau^5^, with a canonical KFERQ sequence or with posttranslational modifications that mimic the KFERQ motif (e.g. phosphorylation, acetylation) ^38, 39, 40, 41^. To determine whether *FAM151B-DT* interacts with other proteins with KFERQ and KFERQ-like sequences, we used a silico database that has defined KFERQ and KFERQ-like sequences (https://rshine.einsteinmed.edu/) ^38^. We found that 81% of the *FAM151B-DT* proteome contained KFERQ-like motifs (**Figure 6A**; **Supplemental Table 11**). These motifs were categorized based on their occurrence, with canonical motifs being the most prevalent, followed by phosphorylation- and acetylation-generated motifs (**Figure 6A**; **Supplemental Table 11**). Together, this finding suggests that *FAM151B-DT* may work cooperatively with HSC70 to target and deliver proteins for degradation in the lysosome.

**Figure 6.**
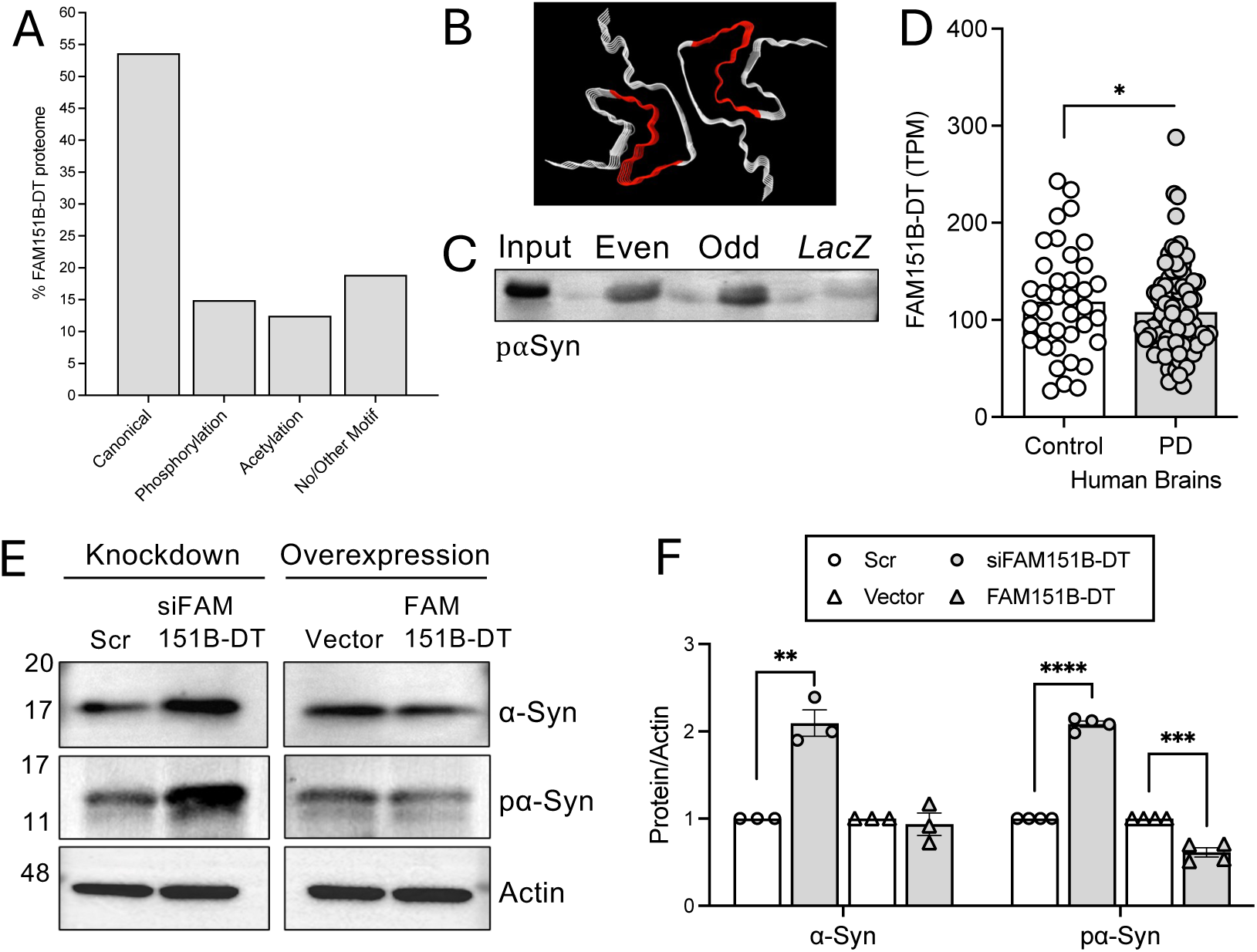
*FAM151B-DT* interacts with autophagy substrates implicated in Parkinson’s disease. A. The presence of KFERQ-like motifs were evaluated among the *FAM151B-DT* interactome (n=101). Motif types were plotted. B. Crystal structure of alpha-synuclein (PDB: 6a6b, ^53^) illustrating the FAM151B-DT binding sites in red based on ChIRP-MS. C. ChIRP-immunoblot of *FAM151B-DT* pull-down fractions (see Figure 4E) probed with pα-Syn antibody in SH-SY5Y cells. D. *FAM151B-DT* expression is significantly reduced in PD brains compared with neuropathology free controls. *, p=0.04. E. SH-SY5Y cells were transiently transfected with scrambled (scr) or *siFAM151B-DT* siRNAs and control vector or FAM151B-DT overexpression plasmid. Representative western blots of phosphorylated (Ser199, pα-Syn) and total α-synuclein (α-Syn). F. Quantification of western blot data from (E). *FAM151B-DT* silencing: white circle, scrambled (scr); gray circle, *siFAM151B-DT*. *FAM151B-DT* overexpression: white arrow, vector; gray arrow, *FAM151B-DT*. Data are presented as mean ± SEM from four independent experiments. Student’s *t*-test: **, p<0.01; ***, p<0.001; ****, p<0.0001.

The α-synuclein protein, which accumulates into Lewy bodies in Parkinson’s disease, contains a KFERQ-like motif (VKKDQ) ^42, 43, 44^. Tau and α-synuclein share many other functional properties including roles in synaptic activity ^20, 45, 46, 47^, secretion via the unconventional secretory pathway ^48, 49^, and seeding and spreading of protein aggregates in a prion-like manner ^50, 51, 52^. To determine whether *FAM151B-DT* interacts with α-synuclein, we analyzed our ChIRP-MS dataset. Using a relaxed stringency (total spectral count ≥ 1, ≥90% peptide thresholds, and ≥80% protein thresholds; **Supplemental Table 12**), we discovered α-synuclein. The interaction sites were mapped on to a representative crystal structure of α-synuclein (**Figure 6B**; PDB: 6a6b; ^53^). RNA-IP of *FAM151B-DT* in SH-SY5Y cells followed by immunoblotting of α-synuclein further validated our mass spectrometry findings (**Figure 6C**). Thus, *FAM151B-DT* interacts with α-synuclein.

Our interest in *FAM151B-DT* began with a finding that its expression was reduced in *MAPT* mutant neurons and tauopathy brains (**Figure 1**). Our finding that like tau, *FAM151B-DT* interacts with α-synuclein led us to consider whether FAM151B-DT expression is also altered in brains from Parkinson’s disease (PD) patients. Bulk RNA sequencing data revealed a significant reduction of *FAM151B-DT* levels in PD brains compared with neuropathology free control brains (p=0.04; **Figure 6D**). Thus, *FAM151B-DT* expression is altered in a similar pattern in FTLD-tau, PSP, AD, and PD brains.

Given that *FAM151B-DT* expression regulates autophagy and alters ptau and total tau levels in SH-SY5Y cells, we sought to determine how α-synuclein is impacted by modifying *FAM151B-DT* levels. Silencing *FAM151B-DT* in SH-SY5Y cells resulted in a significant increase in phosphorylated α-synuclein and total α-synuclein (**Figure 6E-F**). Conversely, overexpressing *FAM151B-DT* led to a significant reduction of phosphorylated α-synuclein without impacting total α-synuclein (**Figure 6E-F**). This pattern is consistent with the impact of *FAM151B-DT* expression on ptau and total tau levels. Together, we demonstrate that *FAM151B-DT* regulates protein homeostasis, including autophagy, in a manner that can impact tau and α-synuclein accumulation.

## Discussion

Here, we discovered a novel lncRNA, *FAM151B-DT*, that is altered in a stem cell model of tauopathy and in brains from numerous neurodegenerative diseases, including FTLD-tau, PSP, AD, and PD. By mimicking the expression pattern of *FAM151B-DT* that we observed in stem cell models and brain tissue, we found that *FAM151B-DT* silencing promotes tau aggregation *in vitro*. We demonstrate that *FAM151B-DT* interacts with tau, α-synuclein, the chaperone HSC70, and a number of other proteins enriched in protein homeostasis. Silencing *FAM151B-DT* expression stymies autophagic flux, leading to accumulation of p62 as well as proteins that are prone to aggregate ptau, total tau, p-α-synuclein, and α-synuclein. We go on to show that restoring *FAM151B-DT* expression is sufficient to enhance autophagic flux, lower ptau and p-α-synuclein, and reduce tau aggregation. Thus, *FAM151B-DT* is a novel therapeutic target that could benefit several neurodegenerative diseases.

We discovered that *FAM151B-DT* is a poorly conserved cytoplasmic lncRNA. ChIRP profiling of the *FAM151B-DT* interactors point to functions involved in maintaining cellular structure and functions ^54 55^ including protein processing, stress response, and RNA transport. Our findings reveal a context-dependent role of *FAM151B-DT* in regulating autophagy pathways, with significant implications for cellular protein homeostasis and the degradation of pathogenic proteins, such as phosphorylated tau ^5^.

Mutations in the *MAPT* gene, such as P301L, IVS10+16, and R406W, are sufficient to cause familial forms of FTLD-tau ^28^. These mutations induce structural changes in the tau protein, promoting hyperphosphorylation, misfolding, and aggregation into NFTs, a hallmark of tauopathies, including FTD, PSP, and AD ^56, 57^. Tau clearance depends on efficient protein degradation pathways, including autophagy ^58^. In macroautophagy, autophagosomes sequester tau aggregates, deliver them to lysosomes, and facilitate their degradation ^6^. Tauopathies impair this process through lysosomal dysfunction, defects in autophagosome formation, or reduced autophagic flux ^28^. Additionally, tau can suppress autophagy by activating the mTOR pathway, which inhibits autophagy initiation ^32^. Restoring autophagy can be achieved through AMPK activation, which suppresses mTOR and enhances autophagic flux. In chaperone mediated autophagy, tau proteins containing KFERQ-like motifs are selectively bound by HSC70, delivered to lysosomes through LAMP2A, and subsequently degraded ^6^. However, hyperphosphorylated tau blocks CMA clearance and reduces LAMP2A levels, further impairing CMA and accelerating tau pathology ^5^.

Our findings reveal that *FAM151B-DT* regulates autophagy pathways. Silencing of *FAM151B-DT* impairs autophagic flux, likely at the lysosomal degradation stage, resulting in inefficient clearance of pathogenic proteins, an underlying feature of neurodegeneration. Conversely, overexpression of *FAM151B-DT* enhances autophagy, facilitating the selective degradation of phosphorylated tau and reducing its accumulation. LncRNAs have recently emerged as critical regulators of gene expression and cellular functions ^8^. Among these roles, lncRNAs have drawn interest for their ability to regulate autophagy which is a vital cellular process responsible for degrading and recycling damaged organelles, misfolded proteins, and aggregates such as tau. Emerging evidence shows that lncRNAs can directly interact with autophagy components or modulate autophagy-related signaling pathways ^9, 59^. Specific lncRNAs have been demonstrated to activate autophagy, thereby mitigating pathological protein accumulation and providing neuroprotective effects ^60, 61, 62^. Moreover, lncRNAs influence autophagy by regulating key components such as LC3, p62, and lysosomal proteins, thereby controlling both the initiation and completion of autophagic flux ^63, 64, 65^. This regulatory interplay provides a powerful mechanism to clear toxic aggregates, alleviate cellular stress, and slow neurodegeneration, offering promising therapeutic avenues for tauopathies and related disorders. Given their diverse regulatory functions, lncRNAs hold significant therapeutic potential, particularly for targeting autophagy. Enhancing protective lncRNA expression or mimicking their functions could restore autophagic balance, clear pathological protein aggregates, and alleviate disease symptoms.

Our discovery of an interaction between *FAM151B-DT* and HSC70 and tau supports a role for *FAM151B-DT* in maintaining proteostasis. HSC70, a constitutively expressed member of the HSP70 molecular chaperone family in mammals, plays a fundamental role in maintaining proteostasis under both physiological and stress conditions ^66^. It interacts with various substrates, including newly synthesized polypeptides, folding intermediates, misfolded proteins, and aggregates, while also assisting in uncoating clathrin-coated vesicles ^67^. By binding to exposed hydrophobic regions, HSC70 prevents misfolded proteins from aggregating and facilitates their degradation via the ubiquitin-proteasome system and autophagy pathways ^67^. HSC70 helps solubilize protein aggregates through ATP-driven mechanisms in conjunction with co-chaperones ^37^. HSC70 is pivotal to the process of macroautophagy, chaperone mediated autophagy, and endosomal micro-autophagy ^68 69, 70 71, 72, 73^. HSC70 interacts with tau to facilitate proper folding and to prevent the accumulation of misfolded tau, which is implicated in NFT formation in AD ^74^. HSC70 mediates the degradation of unmodified tau and α-synuclein through chaperone mediated autophagy, endosomal micro-autophagy and ubiquitin proteosome pathways ^25, 69, 74^.

We show that *FAM151B-DT* plays a key role in regulating autophagy via HSC70. Based on this finding, we hypothesized that *FAM151B-DT* might serve as a scaffold to help support positioning of client proteins with HSC70 to facilitate proper folding and/or targeting to the lysosome for degradation. Consistent with this hypothesis, we found that more than 80% of *FAM151B-DT* interactors contain HSC70 recognition sequences that are essential for several forms of autophagy, including chaperone mediated autophagy. Other HSC70 client proteins that are implicated in neurodegenerative diseases, like α-synuclein, are also affected by *FAM151B-DT*. A well-recognized chaperone mediated autophagy substrate, α-synuclein, undergoes degradation via chaperone mediated autophagy under physiological conditions^44^. Tau and α-synuclein directly interact with LAMP2A, a critical receptor for CMA activity^5, 42, 75^. Targeting HSC70, via miR-320a, suppresses activity and promotes protein accumulation^76^. With age, proteostasis decreases, including autophagy, and this is exaggerated in neurodegenerative disease.

In conclusion, we discovered a novel lncRNA, *FAM151B-DT*, that plays a pivotal role in autophagy regulation. We show that *FAM151B-DT* facilitates the degradation of aggregation prone proteins associated with neurodegenerative diseases such as tau and α-synuclein. Promoting *FAM151B-DT* expression or enhancing its interaction with HSC70 may represent a therapeutic opportunity to restore efficient protein degradation, reduce pathological protein accumulation, and counteract age-related autophagy decline, offering promising avenues for neurodegenerative disease intervention.

## Methods

### Patient consent

The informed consent was approved by the Washington University School of Medicine Institutional Review Board and Ethics Committee (IRB 201104178 and 201306108). The University of California San Francisco Institutional Review Board approved the operating protocols of the UCSF Neurodegenerative Disease Brain Bank (from which brain tissues were obtained). Participants or their surrogates provided consent for autopsy, in keeping with the guidelines put forth in the Declaration of Helsinki, by signing the hospital’s autopsy form. If the participant had not provided future consent before death, the DPOA or next of kin provided it after death. All data were analyzed anonymously.

### iPSC generation and genome engineering

Human iPSCs used in this study have been previously described ^77^. iPSC lines were generated using non-integrating Sendai virus carrying the Yamanaka factors: OCT3/4, SOX2, KLF4, and cMYC (Life Technologies) ^78, 79^. The following parameters were used for the characterization of each of the iPSC lines using standard methods ^77^: pluripotency markers by immunocytochemistry (ICC) and quantitative PCR (qPCR); spontaneous or TriDiff differentiation into the three germ layers by ICC and qPCR; assessment of chromosomal abnormalities by karyotyping; and *MAPT* mutation status confirmation by Sanger sequencing (characterization data previously reported ^80^).

To determine the impact of the *MAPT* mutant allele on molecular phenotypes, we used CRISPR/Cas9-edited isogenic controls in which the mutant allele was reverted to the wild-type (WT) allele in each of the donor iPSC lines as previously described ^77, 80^. The resulting edited iPSC lines were characterized as described above in addition to on- and off-target sequencing (characterization data previously reported ^80^). All iPSC lines used in this study carry the *MAPT* H1/H1 common haplotype. All cell lines were confirmed to be free of mycoplasma.

### Differentiation of iPSCs into cortical neurons

iPSCs were differentiated into cortical neurons as previously described ^4, 77, 80, 81^(https://dx.doi.org/10.17504/protocols.io.p9kdr4w). Briefly, iPSCs were plated at a density of 65,000 cells per well in neural induction media (StemCell Technologies) in a 96-well v-bottom plate to form neural aggregates. After 5 days, cells were transferred into culture plates. The resulting neural rosettes were isolated by enzymatic selection (Neural Rosette Selection Reagent; StemCell Technologies) and cultured as neural progenitor cells (NPCs). NPCs were differentiated in planar culture in neuronal maturation medium (neurobasal medium supplemented with B27, GDNF, BDNF, and cAMP). The cells were analyzed after 6 weeks in neuronal maturation medium. Currently, tau protein levels are stable and similar to protein profiles described in human brains ^82^.

### RNA sequencing and differential expression analyses

RNAseq was generated from iPSC-derived neurons as previously described ^80, 81,11^. Briefly, samples were sequenced by an Illumina HiSeq 4000 Systems Technology with a read length of 1×150 bp and an average library size of 36.5 ± 12.2 million reads per sample. Salmon (v. 0.11.3) ^83^ was used to quantify the expression of the genes annotated within the human reference genome (GRCh38.p13). The lncRNA genes were selected for downstream analyses. LncRNA genes that were present in at least 10% of samples with expression >0.1 TPM were included in subsequent analyses: 7,537 lncRNA genes^11^. Principal component analyses (PCA) were performed with the selected 7,537 non-coding genes using regularized-logarithm transformation (rlog) counts. Differential gene expression was performed using the DESeq2 (v.1.22.2) R package ^84^. PCA and differential gene expression analyses were performed independently for each pair of *MAPT* mutations and isogenic controls. Each *MAPT* mutation and its isogenic control were considered independent cohorts due to their shared genetic background. PCA and Volcano plots were created for each comparison using the ggplot2 R package (v3.3.6) ^85^.

### Gene expression analysis in PSP and AD brains

To determine whether the 15 common differentially expressed lncRNAs identified in the *MAPT* mutant iPSC derived neurons were altered in brains from primary tauopathy patients, we analyzed gene expression in a publicly available dataset: the temporal cortex of 76 control, 82 PSP, and 84 AD brains (syn6090813) ^86^. Differential gene expression analyses comparing controls with PSP and AD brains were performed using a “Simple Model” that employs multi-variable linear regression analyses using normalized gene expression measures and corrected by sex, age-at-death, RNA integrity number (RIN), brain tissue source, and flowcell as covariates ^86^. Transcriptomic data from the middle temporal gyrus of FTLD-tau patients with *MAPT* IVS10+16 and p.P301L mutation (*MAPT* IVS10+16 n=2 and *MAPT* p.P301L n=1) and neuropathology free controls (n=3) were also analyzed ^11, 80^. Differential expression analyses comparing FTLD-tau mutation carrier brains with controls were performed using DESeq2 (v.1.22.2) R package ^84^ as previously described ^11, 80^.

### Gene expression analysis in PD brains

Bulk RNA sequencing was performed in samples from the parietal lobe of subjects with PD (n=99) and neuropathology free controls (n=38). Research subjects were assessed in the Movement Disorders Center and the Knight Alzheimer Disease Research Center at Washington University in St. Louis (WUSTL). All clinical information was obtained from the brain bank database and neuropathologic examinations were performed using the WUSTL protocol ^87, 88^. A neuropathology diagnosis of PD was based on the presence of Lewy bodies, Lewy neuritis, and hypopigmentation of the substantia nigra. Brain sample preparation and quality control measures were performed as previously described ^89^. Samples that were historically processed in three different databases; thus, we used ComBat-Seq (sva package, v3.52.0) ^90^ to minimize the batch effect. DESeq2 (v1.44.0) ^84^ was then employed to generate the differential gene expression. Samples with normalized counts outside two standard deviations in the principal component analysis were excluded. The model compared expression patterns of PD cases with controls (status) and included sex, age at death, median TIN, and databases as covariates (model ∼ sex + AgeDeath + TIN_median + DatabaseA + DatabaseB + status). Databases were entered as dummy covariates. FDR was used to adjust the *p*-value for multiple test correction.

### Cell Culture

Human SH-SY5Y and HEK293T tau Biosensor cells ^12^ were cultured in Dulbecco’s Modified Eagle Medium (DMEM; 61965–059, Gibco), supplemented with 10% fetal bovine serum (FBS; 10270106, Gibco), 1% non-essential amino acids (11140035, Gibco), and 1% penicillin-streptomycin (15140122, Gibco). Cells were maintained in a humidified atmosphere with 5% CO₂ at 37°C and were used within 5–10 passages.

### Transient Transfection in Immortalized Cell Lines

siRNAs targeting *FAM151B-DT* were designed and synthesized by Thermo Fisher Scientific. The sequences were as follows: siRNA1 Sense: 5’-GCUCACAUGUUGUCCAACUTT-3’; Anti-sense: 5’-AGUUGGACAACAUGUGAGCTT-3’ and siRNA2 Sense: 5’-CCUGGUGCUUCCUGUAAAUTT-3’; Anti-sense: 5’-AUUUACAGGAAGCACCAGGTT-3’. SH-SY5Y or HEK293T tau Biosensor cells were transfected with siRNA using Lipofectamine RNAiMAX (Invitrogen) following the manufacturer’s instructions. Briefly, cells were seeded in a 6- or 24-well plate at 60-70% confluence. 100 nM of siRNA was mixed with 5 μl of Lipofectamine RNAiMAX in 500 μl serum-free Opti-MEM (Gibco, USA) and incubated at room temperature for 20 minutes to form complexes. After washing cells with PBS, the 500 μl transfection mixture was added to each well. After 24 hours, the medium was replaced with fresh DMEM containing 10% FBS. Cells were harvested 48 hours post-transfection for downstream analysis.

For transient transfection of plasmids, cells were grown in DMEM supplemented with 10% FBS, 1% L-glutamine, and 1% penicillin-streptomycin. The full-length cDNA of FAM1515B-DT was synthesized by Genscript and cloned into the pCDNA3.1^+^ expression vector along with tdTomato reporter. Control plasmids and plasmid-mediated FAM151B-DT overexpression, were obtained from Genscript (Genscript, USA). pcDNA3.1+-FAM151B-DT-tDT or control pcDNA3.1+-tDT plasmid DNA constructs (Genscript, USA) were transfected using Lipofectamine 2000 (Invitrogen, San Diego, CA, USA) according to the manufacturer’s protocol. The transfected cells were evaluated after 48 hours for further analysis.

### Comprehensive identification of RNA-binding proteins by mass spectrometry (ChIRP-MS) Assay

ChIRP-MS was carried out in SH-SY5Y cells as previously described^18^. *FAM151B-DT* specific anti-sense probes with BiotinTEG at 31 end were designed using the Biosearch Technologies ChIRP Probe Designer tool (https://www.biosearchtech.com/chirpdesigner/) with maximum repeat masking setting and even coverage of the whole transcript (**Supplemental Table 2**). The LacZ probe with BiotinTEG at its 31 end was used as a negative control. Briefly, SH-SY5Y cells were crosslinked with 1% formaldehyde and lysed using Lysis Buffer (50 mM Tris-HCl [pH 7], 10 mM EDTA, 1% SDS). The cell extract was sonicated in a Qsonic Bath sonicator for 10 cycles (301 ON/451 OFF) at 50% amplitude and centrifuged at 16,000xg for 10 minutes at 4°C to eliminate insoluble chromatin. One percent of the cleared extract was reserved as input, while the remaining material was diluted with Hybridization Buffer (15% formamide, 500 mM NaCl, 1 mM EDTA, 0.5% SDS, supplemented with protease and RNase inhibitors), incubated with biotinylated *FAM151B-DT* even and odd oligos, and rotated at 37°C overnight in a hybridization chamber. A pool of five different biotinylated odd and even-positioned oligonucleotides was used to pull down *FAM151B-DT*, with four oligonucleotides recognizing the LacZ gene as independent controls. A total of 100 pmol of probes was used per reaction. The following day, 100 µl of equilibrated Streptavidin magnetic beads (Dynabeads MyOne Streptavidin C1 - Thermo Fisher) were added to each pull-down condition and incubated for 2 hours at 37°C in a hybridization chamber. Beads were washed five times with Wash Buffer (2× saline-sodium citrate [SSC], 0.5% SDS), and elution was performed using Protein Kinase buffer (100 mM NaCl, 1 mM EDTA, 0.5% SDS, 10 mM Tris-HCl [pH 7] or [pH 8] for RNA or DNA, respectively). Ten percent of the washed beads were used to analyze RNA enrichment by one-step qPCR after standard RNA extraction, while the remaining beads were subjected to on-bead digestion followed by mass spectrometry analysis.

### Mass spectrometry

#### On-bead digestion

Trypsin (800 ng) was added to immunoprecipitated proteins on the beads for 6 hours at 37°C. The initial digested samples were centrifuged for 2 minutes at 5,000 x g and the supernatants were collected into fresh tubes. Beads were washed twice with 100 mM ammonium bicarbonate and the supernatants were pooled. The resulting samples were reduced with 20 mM dithiothreitol at 37°C for 1 hour, and cysteine was alkylated with 80 mM iodoacetamide for 45 minutes in dark. Samples were treated with 600 ng of trypsin to overnight incubation at 37°C. The resulting peptides were desalted using solid-phase extraction on a C18 Spin column and eluted with 0.1% FA in 80% ACN.

#### LC-MS and data analysis

Peptides were injected onto a Neo trap cartridge (Thermo Fisher Scientific) coupled with an Aurora Ultimate TS 25 cm analytical column (IonOpticks). Samples were separated using a linear gradient of solvent A (0.1% formic acid in water) and solvent B (0.1% formic acid in ACN) over 120 minutes using a Vanquish Neo UHPLC System coupled to an Orbitrap Ascend Tribrid Mass Spectrometer (Thermo Fisher Scientific). The resulting tandem MS data was queried for protein identification against the SwissProt human protein database using Mascot v.2.8.3 (Matrix Science). The following modifications were set as search parameters: peptide mass tolerance at 10 ppm, trypsin enzyme, 3 allowed missed cleavage sites, carbamidomethylated cysteine (static modification), and oxidized methionine, deaminated asparagine/glutamine, and protein N-term acetylation (variable modification). The search results were validated with 1% FDR of protein threshold and 90% of peptide threshold using Scaffold v5.3.0 (Proteome Software).

### Tau Seeding Assay

To investigate whether changes in *FAM151B-DT* expression modulate tau seeding activity, we utilized a tau biosensor cell line expressing the tau repeat domain (RD) with the FTD-linked P301S variant of tau fused to either CFP or YFP ^12^. Cells were cultured in 12-well plates and transfected with *FAM151B-DT* siRNA, overexpression plasmids, control siRNA, or control plasmids. After 24 hours, 10 µM of human recombinant Tau-441 (2N4R) wild-type protein pre-formed fibrils (StressMarq, USA) were transfected using 5 µL of Lipofectamine 2000. Following 48 hours, cells were harvested with 0.05% trypsin, post-fixed in 4% paraformaldehyde (Electron Microscopy Services) for 5 minutes, and resuspended in flow cytometry buffer (2% FBS, 0.5% BSA in PBS). FRET flow cytometry was performed using a BD LSR Fortessa™ Cell Analyzer (BD Biosciences). CFP and FRET were measured by exciting cells with a 405 nm laser, with fluorescence captured at 405/50 nm and 525/50 nm, respectively. YFP was measured by exciting cells with a 488 nm laser, capturing fluorescence at 525/50 nm. FRET signal was quantified following Banning et al. ^91^, with cells expressing CFP or YFP alone used to compensate for signal overlap. A final bivariate plot of FRET vs. CFP was generated with a triangular gate to assess FRET-positive cells. This gate was adjusted using biosensor cells treated with lipofectamine alone as FRET-negative controls and Zombie Aqua positive cells as FRET-positive controls. For each experiment, 50,000 cells per replicate were analyzed, with data processed using FlowJo v10 software.

### RNA Immunoprecipitation

To measure the molecular interaction between tau, HSC70, and *FAM151B-DT*, RNA immunoprecipitation was performed as previously described with minor modifications ^92^. Briefly, SH-SY5Y cells were grown in a T-75 cm² flask to approximately 90% confluency, then trypsinized and pelleted at 300 × g for 5 minutes at room temperature. Cell pellets were washed once with 1X PBS and resuspended in 1 mL of ice-cold lysis/RIP buffer containing 20 mM Tris-HCl (pH 8.0), 200 mM NaCl, 1 mM EDTA, 1 mM EGTA, 0.5% Triton X-100, 0.4 U/µL RNase inhibitor, and 1X protease inhibitor cocktail (Roche). The samples were mixed for 30 minutes at 4°C in an end-over-end rotor. The pelleted samples were sonicated in a Qsonic bath sonicator for 5 cycles of 10 seconds ON and 30 seconds OFF at 50% amplitude to obtain whole cell lysate. The lysates were cleared by centrifugation at 10,000 × g for 15 minutes at 4°C to remove insoluble debris. The supernatant was collected in a separate tube, and the total protein concentration was measured using a BCA assay (Pierce) kit. Approximately 1 mg of pre-cleared lysate was incubated with 2.5 µg of each anti-HSC70 (Invitrogen, MA3-014), Tau5 antibodies (a generous gift from the Binder lab), or pre-immune IgG (sc-2025) overnight at 4°C. The lncRNA-protein complexes were captured with antibody-coupled Protein A/G beads (Thermo Fisher Scientific, Cat#20333), washed with RIP buffer, and treated with RNase-free DNase I. RNAs were isolated from 20% of the beads using the Trizol method. qPCR was performed with FAM151B-DT and GAPDH primers using the iTaq One-Step RT-PCR kit (Bio-Rad).

For western blots, the remaining beads were washed three times with cell lysis buffer and once with 1X phosphate-buffered saline (PBS). The washed beads were mixed and boiled in SDS sample buffer containing β-mercaptoethanol (Bio-Rad), resolved by SDS-PAGE, and transferred onto a PVDF membrane (Immobilon). Membranes were blocked with 5% nonfat dry milk (or 3% BSA) and incubated with primary antibodies, followed by three washes with 0.1% PBS-Tween 20. Blots were then incubated with horseradish peroxidase-conjugated secondary antibodies, followed by another three washes. Signals were visualized using a chemiluminescent detection system, and images were captured with a ChemiDoc Imaging System (Bio-Rad). All blots represent at least three independent experiments, with Actin probed as the loading control.

### Immunoblotting Analysis

Experimental cells were harvested by trypsinization, washed with ice-cold 1X PBS containing protease inhibitors, and then centrifuged at 8000xg for 5 minutes at 4°C. The cell pellets were stored at −80°C until use. Frozen pellets were lysed by sonication in RIPA buffer containing 50 mM Tris (pH 7.4), 150 mM NaCl, 1% Triton X-100, 1% sodium deoxycholate, 0.1% SDS, and inhibitors (50 mM NaF, 1 mM Na2VO4, EDTA, phosphatase, and protease inhibitors from Millipore-Sigma). Lysates were cleared by centrifugation and protein concentration was determined using the BCA assay (Pierce). Equal amounts of protein (10-20 µg) were boiled in SDS sample buffer containing β-mercaptoethanol (Bio-Rad), resolved by SDS-PAGE, and transferred onto a PVDF membrane (Immobilon). Membranes were blocked with 5% nonfat dry milk (or 3% BSA) and incubated with primary antibodies (**Supplemental Table 13**), followed by three washes with 0.1% PBS-Tween 20. Blots were then incubated with horseradish peroxidase-conjugated secondary antibodies, followed by another three washes. Signals were visualized using a chemiluminescent detection system, and images were captured with a ChemiDoc Imaging System (Bio-Rad). All blots represent at least three independent experiments, with Actin probed as the loading control.

### Quantitative PCR (qPCR)

RNA expression was analyzed by real-time PCR. FAM151B-DT was measured using SYBR Green iTaq Universal SYBR Green Supermix (Bio-Rad, USA; **Supplemental Table 14**). *TFEB*, *LAMP2*, *CTSD,* and *MAPT* RNA expression were measured using Applied Biosystems TaqMan Gene Expression Assays along with TaqMan Fast Universal PCR Master Mix (**Supplemental Table 14**). Samples were run in duplicate in each plate. To avoid amplification interference, expression assays were run in separate wells from the housekeeping gene *GAPDH*. Real time data was analyzed by the comparative C_T_ method. Average C_T_ values for each sample were normalized to the average C_T_ values for the housekeeping gene GAPDH. The resulting value was then corrected for assay efficiency. Samples with a standard error of 20% or less were re-analyzed.

### Statistical Analysis

For all statistical analyses, an unpaired, Student’s t-test was used to compare differences between two groups, and a one-way ANOVA was used for comparisons among three or more groups using GraphPad PRISM 10 software. A p-value of less than 0.05 was considered statistically significant.

## Data Availability

All data produced in the present study are available upon reasonable request to the authors.

## Acknowledgements

We thank the research subjects and their families who generously participated in this study. We thank Dr. Binder who generously provided the Tau5 antibodies. We thank Torri Ball for thoughtful discussions. Mass Spectrometry analyses were performed by the Mass Spectrometry Technology Access Center at the McDonnell Genome Institute (MTAC@MGI) at Washington University School of Medicine, supported by the Diabetes Research Center/NIH grant P30 DK020579, Institute of Clinical and Translational Sciences/NCATS CTSA award UL1 TR002345, and Siteman Cancer Center/NCI CCSG grant P30 CA091842. This work was supported by access to equipment made possible by the Hope Center for Neurological Disorders, the Neurogenomics and Informatics Center, and the Departments of Neurology and Psychiatry at Washington University School of Medicine. Funding provided by the National Institutes of Health (P30 AG066444, RF1 NS110890, U54 NS123985, K01 AG083215), Rainwater Charitable Organization (CMK), Farrell Family Fund for Alzheimer’s Disease (CMK), and UL1TR002345. Diagrams were generated using BioRender.com.

## Supplemental Figures

**Supplemental Figure 1. Robust expression of *FAM151B-DT* in iPSC-derived neurons and FTLD-tau, PSP, and AD brains.** RNA sequencing and differential gene expression analysis were performed to assess the expression levels of *FAM151B-DT* among all lncRNAs detected after applying a threshold of >0.1 TPM in >20% of the samples. A. Human iPSC-derived neurons from *MAPT* mutations (P301L, IVS10+16, and R406W) compared to their respective isogenic controls. B. FTLD and control brains. C. PSP and control brains. D. AD and control brains. All values are represented as Log10(TPM+1). Red dashed line indicates *FAM151B-DT* expression in each dataset.

**Supplemental Figure 2. Flow analyses of tau seeding in tau biosensor cells where *FAM151B-DT* expression is altered.** HEK293T tau biosensor cells were transiently transfected with scrambled (scr) or *siFAM151B-DT* siRNAs (A) and control vector or *FAM151B-DT* (B). Cells were then treated with tau preformed fibrils (Tau-441). FACS dot plots of Tau-biosensor cells treated with Tau-441.

**Supplemental Figure 3. Evolutionary conservation of human lncRNA-*FAM151B-DT* in mouse and zebrafish.** A. UCSC genome browser showing the base wise conservation of lncRNA-*FAM151B-DT* in 100 vertebrates and 13 mammals. B. UCSC browser indicates that lncRNA-*FAM151B-DT* is poorly conserved among mammals. Middle inset, example of genomic region conserved between human, mouse and zebrafish. Lower inset, non-conserved genomic region.

**Supplemental Figure 4. *FAM151B-DT* expression patterns in human tissues.** A. *FAM151B-DT* RNA levels in normal human tissues from GTEx. Expression in transcripts per kilobase million (TPM). B. *FAM151B-DT* expression in the adult CNS. C. Brain specific different cell types from http://celltypes.org/brain/93.

**Supplemental Figure 5. *FAM151B-DT* does not regulate expression of nearby genes.** A. Schematic representation including UCSC browser view snapshot depicting the cis-located protein coding genes upstream (*FAM151B*) and downstream (*ZFYVE16*) of *FAM151B-DT*. B-D. The impact of *FAM151B-DT* expression in SH-SY5Y cells on proximal protein coding genes, *FAM151B* and *ZFYVE16*, using 2 different siRNAs specific for *FAM151B-DT*. Graphs represent SEM. B. *FAM151B-DT* siRNAs result in a significant reduction in *FAM151B-DT* levels. C. *FAM151B-DT* siRNAs do not alter *FAM151B* levels. D. *FAM151B-DT* siRNAs do not alter *ZFYVE16*. E-F. Expression of *FAM151B* (E) and *ZFYVE16* (F) in human iPSC derived neurons harboring *MAPT* mutations. Data represented as transcripts per million (TPM). G. Heatmap of *FAM151B-DT* and cis-located coding genes levels in human FTLD-tau, AD, PSP and control brain tissues. All values are represented as Log_2_FC. *, p<0.05; **, p<0.01; ***, p<0.001; ****, p<0.0001. ns, not significant.

**Supplemental Figure 6. String network analysis of *FAM151B-DT* proteome reveals four distinct clusters.** A. Conceptual workflow of the *FAM151B-DT* proteome subjected to Cytoscape and pathway enrichment analyses. B. The *FAM151B-DT* proteome was organized into four clusters with a protein-protein interaction enrichment p-value of 1.0e^−16^.

**Supplemental Figure 7. *FAM151B-DT* interactome is altered in *MAPT* mutant neurons and human brains.** A. Upset plot for *FAM151B-DT* interactors that are also significantly differentially expressed in iPSC-derived neurons from *MAPT* mutation carriers compared with isogenic controls. B. Upset plot for *FAM151B-DT* interactors that are also significantly differentially expressed in FTD with tau inclusions, AD, and PSP brains.

